# Preliminary analysis of scRNA sequencing of children’s lung tissues excludes the expression of SARS-CoV-2 entry related genes as the key reason for the milder syndromes of COVID-19 in children

**DOI:** 10.1101/2020.05.25.20110890

**Authors:** Yue Tao, Ruwen Yang, Chen Wen, Jue Fan, Jing Ma, Qiao He, Zhiguang Zhao, Xinyu Song, Hao Chen, Guocheng Shi, Minzhi Yin, Nan Fang, Hao Zhang, Huiwen Chen, Xi Mo

**Affiliations:** The Laboratory of Pediatric Infectious Diseases, Pediatric Translational Medicine Institute, Shanghai Children’s Medical Center, Shanghai Jiao Tong University School of Medicine, Shanghai, China; Department of Cardiothoracic Surgery, Shanghai Children’s Medical Center, Shanghai Jiao Tong University School of Medicine, Shanghai, China; Singleron Biotechnologies, Yaogu Avenue 11, Nanjing, Jiangsu Province, China; Department of Pathology, Shanghai Children’s Medical Center, Shanghai Jiao Tong University School of Medicine, Shanghai, China; Department of Pathology, the Second Affiliated Hospital & Yuying Children’s Hospital of Wenzhou Medical University, Wenzhou, Zhejiang Province, China; Department of Cardiothoracic Surgery, the First Affiliated Hospital of Soochow University, Suzhou, Jiangsu Province, China

**Author notes:** These authors contribute equally to this work (Yue Tao, Ruwen Yang, Chen Wen).

## Abstract

To explore whether the expression levels of viral-entry associated genes might contribute to the milder symptoms in children, we analyzed the expression of these genes in both children and adults’ lung tissues by single cell RNA sequencing (scRNA-seq) and immunohistochemistry (IHC). Both scRNA-seq and IHC analyses showed comparable levels of the key genes for SARS-CoV-2 entry in children and adults, including *ACE2, TMPRSS2* and *FURIN*, suggesting that instead of lower virus intrusion rate, other factors are more likely to be the key reasons for the milder symptoms of SARS-CoV-2 infected children.

## INTRODUCTION

Since December 2019, a novel coronavirus named severe acute respiratory syndrome coronavirus 2 (SARS-CoV-2) has spread rapidly^1^ and posed a severe threat to global public health^2^. As of 31 August 2020, this disease, named coronavirus disease 2019 (COVID-19), had cumulatively caused nearly 25 million confirmed cases and around 800,000 deaths globally, indicating SARS-CoV-2 as a highly contagious virus^3^.

Accumulating data have suggested that the SARS-CoV-2 infection in children is relatively rare and less severe compared to that in adults^4-6^. Meanwhile, recent reports highlight a clinical syndrome in children related to SARS-CoV-2 which is multisystem inflammatory syndrome in children (MIS-C) comprising multiorgan dysfunction and systemic inflammation^7^. However, a recent study pointed out that children are just as likely to be infected by SARS-CoV-2 as adults when exposed to similar environment. Among all children under 10 who had close contact with confirmed cases, the infection rate was 7.4%, which is comparable to that of the whole population average (7.9%)^8^. The latest guidelines also state that all individuals, including children, are generally susceptible to SARS-CoV-2^9^. In the meantime, the low incidence of critical illness in children has been reported repeatedly and accepted widely while the underlying mechanism remains to be elucidated.

Currently there are two major speculations regarding why children display milder symptoms during the infection: relatively lower expression of viral-entry associated genes compared to adults and immune-related factors^10^. Angiotensin converting enzyme 2 (ACE2), a type I membrane protein expressed in various types of organs, has been proven to be the key receptor of SARS-CoV-2 for cellular entry via its interaction with the spike (S) protein^11^ of the virus, which is cleaved by transmembrane protease serine 2 (TMPRSS2)^12^. But it is worth noting that simultaneously blocking the activity of TMPRSS2 and the cysteine proteases CATHEPSIN B/L cannot completely inhibit the entry of SARS-CoV-2 *in vitro*, suggesting the possible involvement of additional proteases in the priming of SARS-CoV-2^12^. A FURIN cleavage site has been identified at S1/S2 boundary in the SARS-CoV-2 S protein^13^, which is suggested to be potentially cleaved by FURIN^14^ as an additional possible mechanism for the priming of the virus. Therefore, in the present study, the expression levels of viral-entry associated genes *(i.e., ACE2, TMPRSS2* and *FURIN)* in both children and adults’ lung tissues were analyzed by single-cell RNA sequencing (scRNA-seq) and immunohistochemistry (IHC) to explore whether the expression levels of these genes might contribute to the milder symptoms in children.

## RESULTS

### Comparable expression of the key genes for SARS-Cov-2 entry by scRNA-seq

Non-affected lung tissues from ten children with congenital heart disease combined with lung diseases requiring lobectomies were collected (Fig. 1A, Supplementary Table S1), and the transcriptome of totally 125,955 cells were analyzed via scRNA-seq. The cells were clustered and annotated into 15 cell types with major cell types known to exist in lung identified (Fig. 1B). When compared to a public dataset (GSE122960) of eight adult lung donors (totally 42,843 cells)^15^, although fewer cells were obtained from children’s lung tissues, the cell subtypes in children were the same as those in adults (Fig. 1B). Consistent with previous analyses^16,17^, AT2 cells (and proliferating AT2 cells) were the major cell type expressing *ACE2*. Other lung/bronchiolar epithelial cells, such as club cells in adults and AT1 cells in children also showed notable expression of *ACE2*. Similarly, the AT2, AT1 and club cells were the major cell types with relatively high expression of *TMPRSS2* in both adults and children. The expression pattern of *FURIN*, however, slightly differed from *ACE2* and *TMPRSS2*, with vascular EC cells and monocyte being the major expressing cell types in adults but AT1 and AT2 cells in children (Fig. 1D). In general, no significant changes were observed in the expression levels of *ACE2, TMPRSS2* and *FURIN* between adults and children, although the percentage of cells expressing *TMPRSS2* and *FURIN* were higher in adults than children (Fig. 1C).

**Fig. 1.**
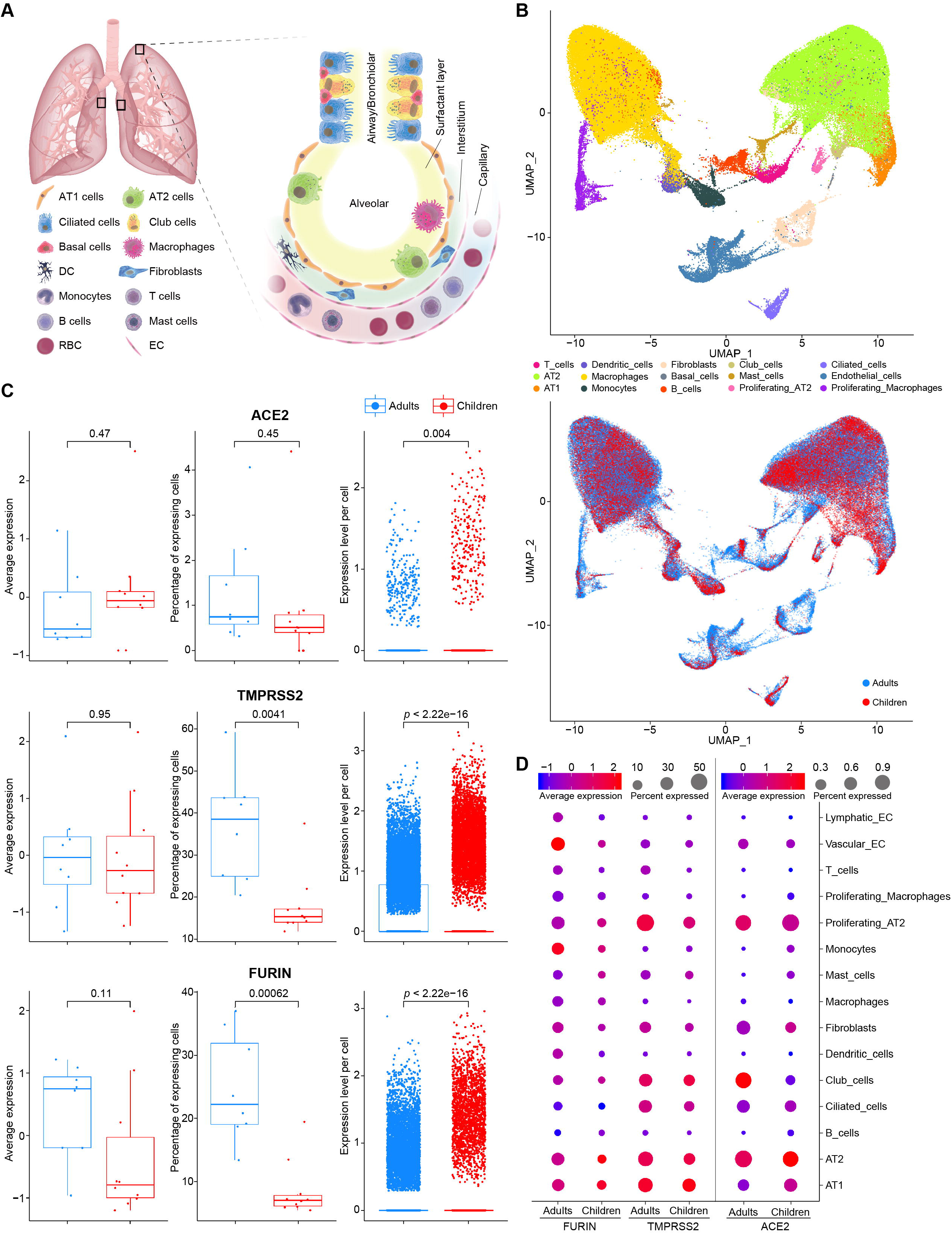
Expression of *ACE2, TMPRSS2* and *FURIN* across different cell types in the lung samples from children and adults determined by scRNA-seq. (**A**) Schematic illustration of the representative sampling locations of the surgical lung specimens from children used in this study and an overview of major cell types detected in the specimens. AT1: alveolar type I; AT2: alveolar type II; DC: dendritic cell; RBC: red blood cell; EC: endothelial cell. Graphic components were obtained and adapted from Library of Science and Medical Illustrations (http://www.somersault1824.com). (**B**) UMAP (Uniform manifold approximation and projection) visualization of an integrated scRNA-seq dataset consists of data from GSE122960 and sequencing results from ten children lung specimens. 15 clustered and annotated cell types are color-coded (upper panel), while cells from adults and children are distinguished with blue dots representing cells from adults and red dots representing cells from children (lower panel). SMC: vascular smooth muscle cell. (**C**) Expression levels of *ACE2, TMPRSS2* and *FURIN* in the lung samples. Average expression level in each sample, percentage of expressing cells in each sample, and expression level of each cell were shown in Box and Whisker Plots (from left to right) and *t*-test was used for statistical analysis. Significance of difference is reflected by the *p*-value shown in the plots. (**D**) Dot-plot illustrating expression of *ACE2, TMPRSS2* and *FURIN* across all cell types in the lung samples. Dot color represents the Z-score normalized average gene expression within each particular cell type. Dot size represents the percentage of cells expressing the respective genes within each cell type. Given the extremely low number of basal cells detected in children’s lung samples, these data points were excluded from the dot-plot for proper visualization of the general view of the gene expression in the majority of the cell types. The complete dataset used for generation of this figure are listed in Supplementary Table S3.

### Comparable expression of the key genes for SARS-Cov-2 entry by IHC

To further verify the expression of *ACE2, TMPRSS2* and *FURIN* at the protein level, IHC was performed in the lung biopsy specimens from children and adults in two independent cohorts (Supplementary Table S2). Consistent with the scRNA-seq analysis, the overall expression levels of ACE2 in children and adults were comparable in both cohorts, with a very small portion of the lung cells expressing ACE2. However, TMPRSS2 and FURIN both showed higher levels in children compared to adults, although the former did not reach statistical significance (Fig. 2A and 2B). The underlying reasons for the partial discrepancy between the results from scRNA-seq and IHC could be the complicated processes downstream of transcription including post-transcriptional, translational and degradation regulations^16^. Meanwhile, the semi-quantitative characteristics of IHC and its limitation in identifying cell types may also contribute to such discrepancies, which further emphasize the necessity to combine these methods for an impartial interpretation of the results. Besides, in accordance with the previous report, the staining patterns of ACE2, TMPRSS2 and FURIN were consistent in the same types of tissue regardless of the pathological condition of the patient^18^.

**Fig. 2.**
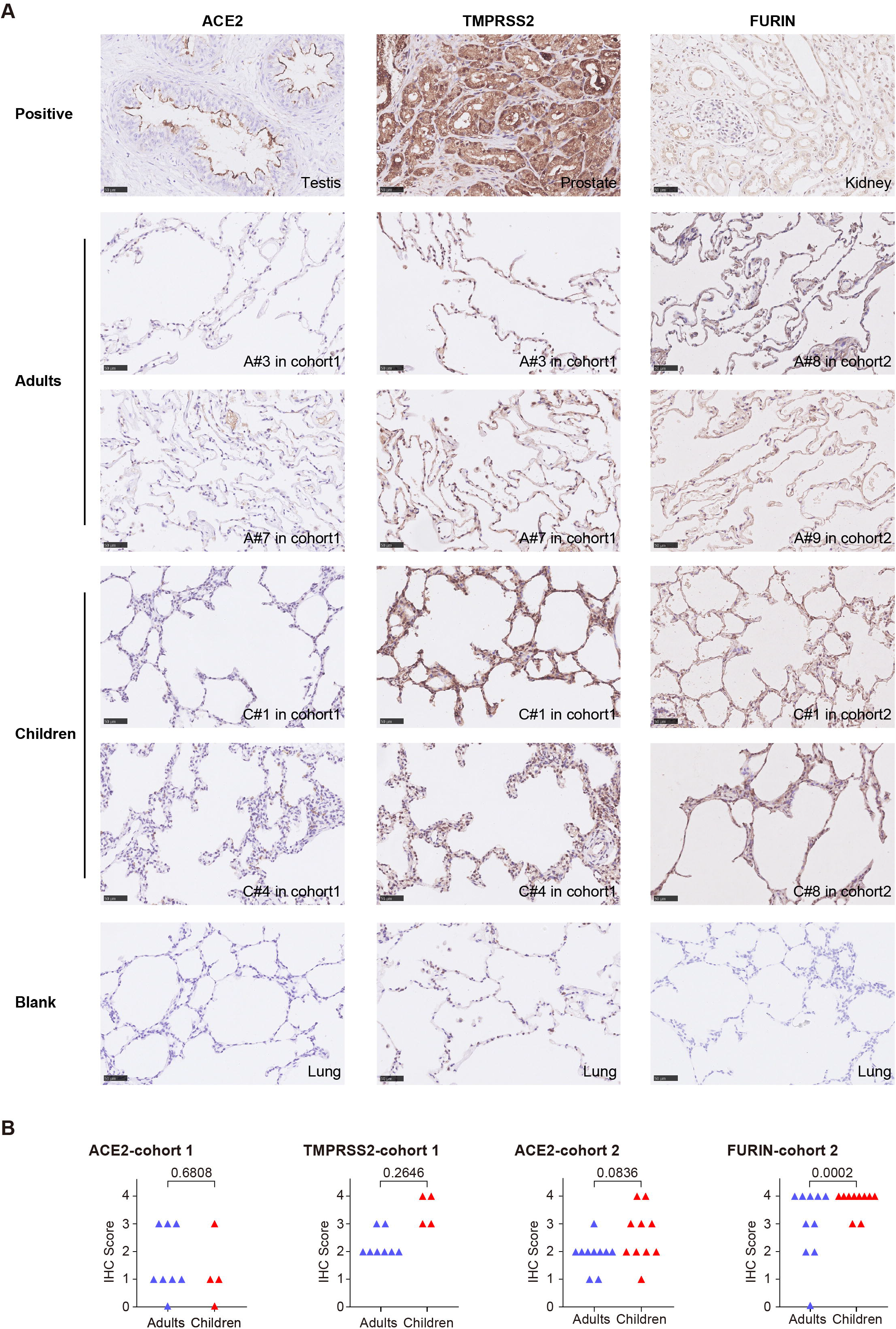
Expression of ACE2, TMPRSS2 and FURIN in the lung samples from children and adults determined by immunohistochemistry (IHC) Sections (4 μm) of formalin-fixed, paraffin-embedded (FFPE) lung tissues from adults and children were subjected to standard IHC protocols to analyze the expression levels of ACE2 and TMPRSS2 (cohort 1) as well as ACE2 and FURIN (cohort 2). Tissues for positive controls of various antibodies are: testis for ACE2, prostate for TMPRSS2 and kidney for FURIN. The graphs are IHC representatives from both cohorts, with the patient number indicated.

## DISCUSSION

Different coronaviruses employ distinct domains within the S1 subunit of S protein to recognize unique entry receptors. SARS-CoV and SARS-CoV-2 interact directly with ACE2 via S domain B to enter the target cells^13,19^, while MERS-CoV utilizes dipeptidyl peptidase 4 (DPP4) as its cellular receptor via S domain A^20^. Both SARS-CoV and MERS-CoV use TMPRSS2 and CATHEPSIN B/L during the priming process. In addition, MERS-CoV S protein can be activated by proprotein convertases and FURIN depending on the host cell and tissue types^21,22^. Although the S proteins of SARS-CoV and SARS-CoV-2 share 76% sequence identity^23^, SARS-CoV-2 S protein harbors a FURIN cleavage site that is absent in SARS-CoV, suggesting its potential cleavage by FURIN^14^.

ACE2 is enriched in the heart, kidney and testis, and is also broadly distributed in the lung, liver, intestine and brain^24^. It is well accepted that the lung is the primary target organ of SARS-CoV-2 infection^25,26^. An analysis of the scRNA-seq data obtained from 43,134 human lung cells showed that *ACE2* was expressed in 0.64 % of all cells in lungs and 83% of the *ACE2* expression was found in AT2 cells, indicating that AT2 is the primary target of SARS-CoV-2 in the lungs^27^, which is consistent with our results. Although children of all ages are susceptible to COVID-19, accumulating clinical data indicated better clinical outcomes in children compared to adults. However, the expression and the activity of ACE2 during the development of children and teenagers are largely unclear^25,28^. Data from animal models remain controversial so far. *ACE2* expression in the lungs was reported to decrease during aging in a study performed in aged rats^29^ and another study using sheep as model revealed significantly lower expression of *ACE2* in the neonatal sheep comparing to that in the adult ones^30^. It has also been shown that higher expression of *ACE2* was observed in the pulmonary alveolar epithelial barrier, cardiomyocytes, and vascular endothelial cells in the aged old non-human primates^31^.

The present study showed comparable expression levels of ACE2, as well as other factors involved in SARS-CoV-2 cellular entry, in children and adults, suggesting that the expression level of viral-entry associated genes is unlikely to be the key reason for milder symptoms in children. Instead, other factors such as unique features in children immunity may play a more important role. Children have a distinct immune profile during infections because their immune system is still under development. It has been shown that compared to adults, decreased mononuclear and polymorphonuclear chemotaxis is found in children and such decrease remains significant until the age of 16^32^. In the meantime, frequent exposure to a plethora of other pathogens, such as respiratory syncytial virus (RSV), would repeatedly challenge the innate immunity including the age dependent maturation of the interferon response^33-36^, possibly leading to an enhanced innate immune function which is one feature of trained immunity^10,37^. Taken together, unique features in children immunity could be a possible explanation for the milder symptoms and lower mortality rate in children SARS-CoV-2 infections.

In summary, the present study, for the first time, described the features of children’s lungs by scRNA-seq. Both scRNA-seq and IHC analyses showed comparable, if not higher, expression of the key genes for SARS-Cov-2 entry, including *ACE2, TMPRSS2* and *FURIN*, suggesting that instead of lower virus intrusion rate, other factors are more likely to be the key reasons for the milder symptoms of SARS-CoV-2 infected children, which awaits further investigation.

## MATERIALS AND METHODS

### Human subjects and specimens

Normal lung tissues from children and adults undergoing biopsies or surgeries for various reasons were collected for scRNA-seq and IHC (Supplementary Table 1). The study was approved by the Institutional Review Board and the Ethics Committee of Shanghai Children’s Medical Center (SCMCIRB-K2020034), and written informed consent was obtained from (the parents of) each patient.

### Tissue dissociation and single cell suspension preparation

Fresh tissue samples were collected and immediately stored in the GEXSCOPETM Tissue Preservation Solution (Singleron Biotechnologies) at 4 °C. Prior to tissue dissociation, the specimens were washed with Hanks Balanced Salt Solution (HBSS) for three times and minced into 1–2 mm pieces. The tissue pieces were digested in 2 ml GEXSCOPE Tissue Dissociation Solution (Singleron Biotechnologies) at 37 °C for 15 min in a 15 ml centrifuge tube with continuous agitation. After digestion, a 40-micron sterile strainer (Corning) was used to separate cells from cell debris and other impurities. The cell suspension was centrifuged at 1000 rpm, 4 °C for 5 min and the cell pellets were resuspended in 1 ml PBS (HyClone). To remove red blood cells, 2 ml GEXSCOPE Red Blood Cell Lysis Buer (Singleron Biotechnologies) was added to the cell suspension and incubated at 25 °C for 10 min. The mixture was then centrifuged at 1000 rpm for 5 min and the cell pellet was resuspended in PBS. The cells were counted with TC20 automated cell counter (Bio-Rad) before further process.

### Single cell RNA sequencing (scRNA-seq) library preparation

Single-cell suspension was adjusted to a concentration of 1×10^5^ cells/ml in PBS, and was then loaded onto a microfluidic chip. The scRNA-seq libraries were constructed according to the manufacturer’s instructions (Singleron GEXSCOPETM Single Cell RNA-seq Library Kit, Singleron Biotechnologies, www.singleronbio.com), and were then sequenced on an Illumina HiSeq X Ten instrument with 150 bp paired end reads. The average sequencing depth per library is around 61 thousand reads per cell.

### Primary analysis of scRNA-seq raw sequencing data

Raw reads were processed to generate gene expression matrices by scopetools (e). Briefly, FastQC (V0.11.7), fastp (1), STAR aligner (2.5.3a) and featureCounts (1.6.2) were used for quality evaluation, trimming, alignment and transcript counting, respectively. Reads without poly(T) tails at the intended positions were filtered out, and cell barcode as well as unique molecular identifier (UMI) for each read were extracted. Adapters and poly(A) tails were trimmed before aligning read 2 to GRCh38 with Ensembl 92 annotation. Reads with the same cell barcode, UMI and gene were grouped together to generate the number of UMIs per gene per cell. Cell number was then determined based on the inflection point of the number of UMI versus sorted cell barcode curve. The scRNA-seq data have been deposited in NCBI’s Gene Expression Omnibus and are accessible through GEO series accession number GSE155900.

### Integrated analysis of children lung data with public adult lung data

The public data of 8 healthy adults were downloaded from Gene Expression Omnibus (GSE122960). Canonical Correlation Analysis (CCA) implemented in Seurat V3 (https://github.com/satijalab/seurat) was used to integrate the adults and children’s datasets. First, cells with less than 200 and more than 5000 genes identified as well as those with less than 30000 UMIs were filtered out. Cells with percentage of mitochondrial genes greater than 20% were also removed. 2000 highly variable genes were used for principal component analysis (PCA) and the first 20 PCs were selected for the subsequent analysis. Resolution 1.25 was used for the clustering analysis to obtain 32 clusters, which were assigned to 15 major cell types based on their canonical marker genes.

### Immunohistochemistry (IHC)

Sections (4 μm) of formalin-fixed, paraffin-embedded (FFPE) lung tissues were subjected to standard IHC protocols to analyze the expression levels of ACE2, TMPRSS2 and FURIN. After deparaffinization, antigen retrieval and blocking, the slides were stained using the Leica Bond (Leica Biosystems) automated staining system with the following primary antibodies for 60 min at room temperature: anti-ACE2 antibody (rabbit, ab108252 from Abcam, 1:100) and anti-TMPRSS2 antibody (rabbit, ab92323 from Abcam, 1:2000) for cohort 1; anti-ACE2 antibody (mouse, 66699-1-Ig, Proteintech, 1:1200) and anti-FURIN antibody (rabbit, 18413-1-AP, Proteintech, 1:1200) for cohort 2. After three washes, the slides were then incubated with HRP-conjugated goat anti-rabbit antibody (cohort 1) or biotin-conjugated anti-mouse/rabbit secondary antibody (cohort 2) for 15-30 min at room temperature. After DAB staining, the slides were counterstained with hematoxylin, dehydrated and mounted. Staining intensities for each antibody were evaluated in a semiquantitative, five-tier manner (negative = 0, partial weak positive = 1, diffused weak positive = 2, partial strong positive = 3, diffused strong positive = 4) by two pathologists independently. The inconsistent scores were determined after consultation by the two pathologists.

### Statistical analysis and data visualization

UMAP (Uniform manifold approximation and projection) visualization of an integrated scRNA-seq dataset consists of data from GSE122960 and sequencing results from ten children lung specimens. 15 clustered and annotated cell types are color-coded, while cells from adults and children are distinguished with blue dots representing cells from adults and red dots representing cells from children. *T*-test was applied to compare the expression levels of *ACE2*, *TMPRSS2* and *FURIN* between children and adults determined by scRNA-seq. The comparison was performed based on library size normalized counts. The normalization was done using NormalizeData function in Seurat V3. The R package ggpubr was used to perform *t*-tests and boxplot visualization. The dot plot was generated using Seurat V3 DotPlot function. Dot color represents the Z-score normalized average gene expression within each particular cell type. Dot size represents the percentage of cells expressing the respective genes within each cell type. Statistical analyses of IHC scores were performed in GraphPad Prism 8 using Mann-Whitney test with an exact *p* value computed. Figures were also generated in GraphPad Prism 8. A *p* value less than 0.05 is considered statistically significant.

## Data Availability

All data can be found in the manuscript and the supplementary materials.

## ACKNOWLEDGMENTS

We would like to thank the patients and their parents for the support and cooperation in publishing this work. We thank the Library of Science and Medical Illustrations (http://www.somersault1824.com) for providing the graphic components used in Fig. 1A.

## AUTHOR CONTRIBUTIONS

Yue Tao, Ruwen Yang, Chen Wen, Hao Zhang, Huiwen Chen and Xi Mo designed the experiment and wrote the paper; Hao Chen and Guocheng Shi provided children’s lung tissues for scRNA-seq; Zhiguang Zhao and Xinyu Song provided the FFPE sections of adults’ lung tissues for IHC; Minzhi Yin provided the FFPE sections of children’s lung tissues for IHC; Qiao He performed IHC; Jing Ma and Minzhi Yin evaluated the IHC score; Ruwen Yang, Jue Fan and Nan Fang analyzed the data from scRNA-seq. All authors participated in the data analysis and manuscript preparation.

